# Study protocol for comparison of post-operative pain between intraosseous multimodal injection and periarticular multimodal injection in Simultaneous Bilateral Total Knee Arthroplasty – A Randomized Controlled Trial

**DOI:** 10.1101/2023.04.29.23289298

**Authors:** Patcharavit Ploynumpon, Vajara Wilairatana, Thakrit Chompoosang

**Affiliations:** Department of orthopedics, Faculty of medicine, Rajavithi hospital, Bangkok, Thailand; Department of orthopedics, Faculty of medicine, Chulalongkorn hospital, Bangkok, Thailand

**Keywords:** Total knee arthroplasty, post operative pain, periarticular infiltration, intra-osseous infiltration

## Abstract

**Background:** Total knee arthroplasty (TKA) was the procedure which is preform by orthopedic surgeons in end stage osteoarthritis knee patients who fail conservative treatment. Periarticular analgesia is one of the effective and safe technique used to reduce pain after surgery. However, the location and formula of the mixture have not yet been controversy. Recently, a study compared intraosseous analgesic injection, which provided significant postoperative pain relief compared to the control group. The authors conducted this study to compare periarticular and intraosseous analgesic injection in patients undergoing simultaneous bilateral total knee arthroplasty.

**Method:** The study is a two-arm, double-blinded, randomized controlled trial with 26 patients (52 knees) who underwent simultaneous bilateral total knee arthroplasty. The primary outcome is postoperative pain at 12, 24, and 48 hours using the Visual Analog Scale for assessment. The secondary outcomes include postoperative range of motion at 24 and 48 hours, postoperative blood loss, and postoperative complications.

**Discussion:** While periarticular analgesic is the standard practice, intraosseous analgesic injection is a potential route for controlling postoperative pain after total knee arthroplasty surgery. This study offers a better alternative route for analgesic injection.

## Background

Total knee arthroplasty (TKA) was the procedure which is preform by orthopedic surgeons in end stage osteoarthritis knee patients who fail conservative treatment. This procedure provide the pain relive and giving back the quality of life to the patients. However the best protocol to optimized the manage after the surgery to reduce the length of hospital stay and reducing post operative pain still on developed in the term Enhanced Recovery After Surgery (ERAS) protocol[1].Multimodal analgesia is one of the mainstay of the pain management in this protocol which are include femoral nerve block, epidural-nerve block, periarticular analgesia, and oral or injected analgesic medication[2].

Periarticular analgesia, which is the injection of the analgesic mixture into surrounding knee tissue by orthopedic surgeon before closing the wound, is one of the effective and safe technique used to reduce pain after surgery. However, the location and formula of the mixture have not yet been controversy.

In the study of Ms. Ava A. Brozovich et al., morphine was injected into the Tibia via intraosseous infusion using a catheter. The study found that the intraosseous group experienced statistically significant reduction in postoperative pain, without an increase in side effects related to morphine use when compared to the placebo group [3].

Currently, there are no studies on injecting the analgesic mixture into intraosseous of the knee. Therefore, we aimed to compare the reduction in postoperative pain between intraosseous infusion and periarticular anesthesia in patients undergoing simultaneous bilateral total knee arthroplasty.

For the safe administration of tranexamic acid, the Drug Therapy Protocols written by the Clinical Quality & Patient Safety Unit (QAS) in Policy code: DTP_TXA_0119, January 2019, provide guidance on the methods of administering tranexamic acid. The protocol states that in adults, tranexamic acid can be safely administered via intraosseous injection (IO) of 1 gram slowly over 2-3 minutes.[4]

The retrospective study of Michael M. Neeki et al. which included 724 trauma patients who had been given 1 gram/10 milliliters of tranexamic acid, was found that the group who were received tranexamic acid had a lower mortality rate compared to the group that did not (3.6% vs. 8.3%; odds ratio = 0.41). The study also shown that tranexamic acid can be given via intraosseous injection in patients who were unable to receive the drug via the intravenous route.[5]

Paoli et al. study found that Ketorolac can be administered via intraosseous injection in truma patients who can not do intravenous admisnisterd.[6]

## Review of the Related Literatures

For the periarticular injection, the surgeon injects the mixture components before suturing the surgical wound at the thigh muscle (quadricep muscle) fascia in front of the knee, joint capsule, collateral ligament. There is no consensus on the formula of the mixture. However, many studies have used a combination regimen containing long-acting analgesia, intravenous NSAID, Tranexamic acid, vasopressor drug [7,8].

The study by Grosu et al. found that local periarticular injection in TKA surgery was safe and simple to perform, and effective in reducing postoperative pain, especially when performed in conjunction with minimal invasive surgery [9]. In addition, periarticular injections were studied compared with control. For example, Fu-Yuen et al.’s study conducted a double-blind crossover RCT comparing multimodal periarticular injection versus femoral nerve block. It found that periarticular injection reduced the postoperative pain score at 72 hours equivalent to femoral nerve block in terms of functional score [10].

A meta-analysis of X.-D. Yun et al.’s study comparing the use of multimodal periarticular injection versus femoral nerve block showed that periarticular analgesia produced a good reduction in postoperative pain at the first 6 hours after surgery more than the nerve block group and produced a similar pain reduction effect up to 48 h (SMD6h = −0.92, 95% CI (−1.38, −0.47))[11]. A systematic review by LinFan et al. compared multimodal periarticular injection and peripheral nerve block; pain scores at rest were significantly lower in the periarticular injection group than in the peripheral nerve block group and were not significantly different [11].

The study by Artit et al. examined the effects of periarthricular injections containing 100 mg bupivacaine, 30 mg, 5 mg morphine sulfate, and 300 mg adrenaline in patients undergoing bilateral knee surgery. The first side was injected in the anterior compartment of the knee, and the other side was injected in the posterior compartment of the knee. It was found that the patient could tell the difference in vas score between both knees, and the injections in the anterior compartment significantly reduced postoperative pain more than injections in the injected posterior compartment of the knee [12].

In the intraosseous injection study, Ava A. et al. conducted a double-blind RCT comparing intraosseous morphine injection into the femoral cavity with a control group. Among TKA patients, postoperative pain medication use was significantly less than the control group in the first 48 hours (40% reduction, P = .001; 49% reduction, P = .036, 38% reduction, P = .025; 33% reduction, P = .041) [3].

However, there is no comparative study of analgesic periarticular injections compared to intraosseous analgesic injections, So the authors conducted this study to compare between periarticular and intraosseous analgesic injection in simultaneous bilateral total knee arthroplasty patients.

## Research question

Does intraosseousinjectionof analgesic mixture result in different postoperative pain control compared to periarticular injection of analgesic mixture in patients undergoing simultaneous bilateral total knee arthroplasty?

### PICO

P Patients scheduled for elective bilateral TKA surgery

I intraosseous injection of analgesia mixture

C periarticular injection of analgesia mixture

O post-operative VAS score, post-operative ROM, rescue analgesia, complication, post operative blood loss

### Objectives

- Primary Objective
  Post-operative VAS score between intraosseous injection and periarticular injection
- Secondary Objective
  Post-operative ROM, rescue analgesia, complication, post operative blood loss

### Hypothesis

H_0_: mean pain score of control group is the same as mean pain score of experimental group

H_1_: mean pain score of control group is different from mean pain score of experimental group

### Conceptual framework

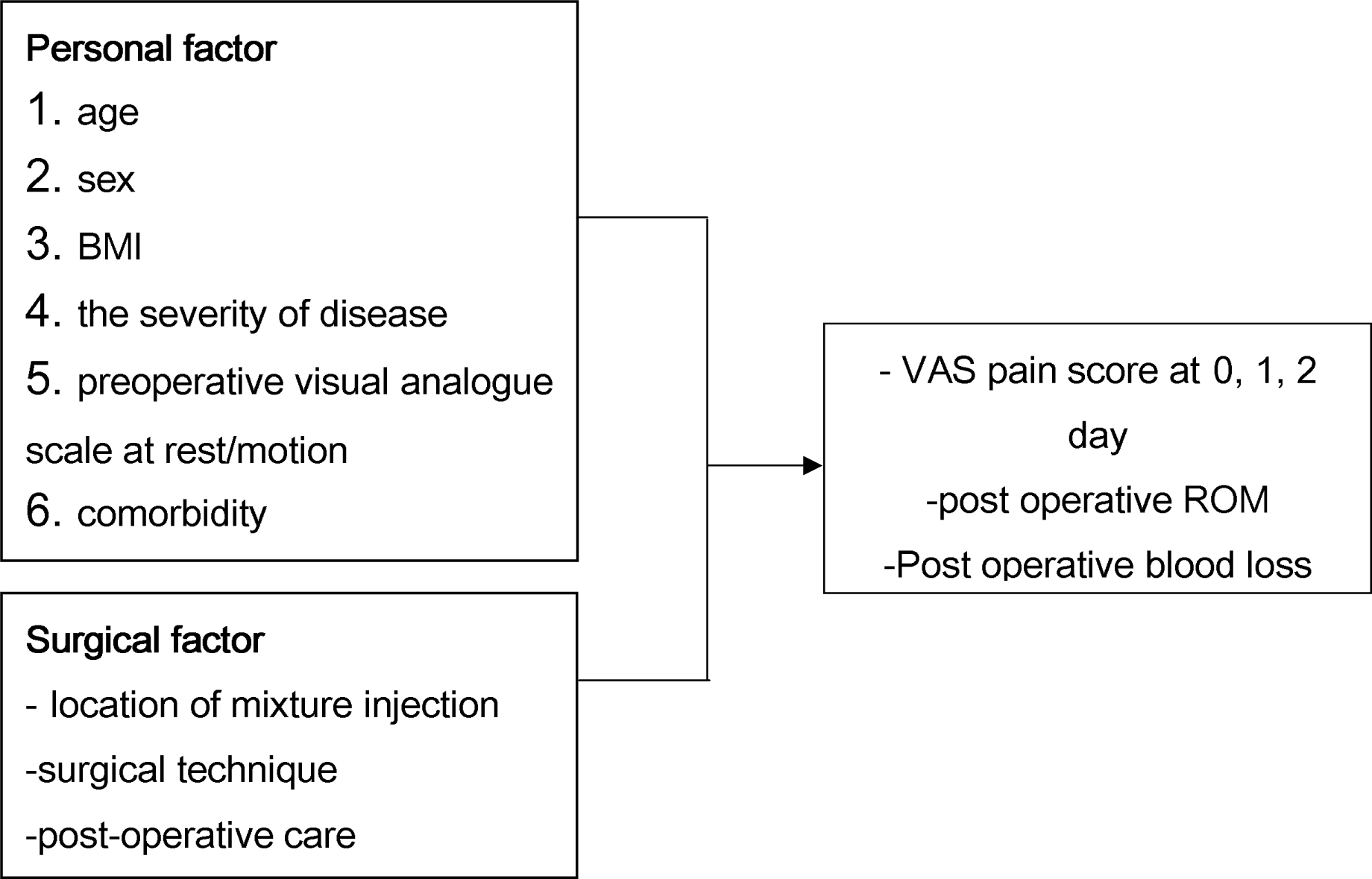

### Operative definition

**Total Knee Arthroplasty:** TKA surgical procedure to replace the distal femoral bone and proximal tibia with special design prothesis

**Length of stay (LOS):** the length of the patients to stay in the hospital after the surgery

**Periarticular multimodal drug injection inject the mixture with comprise of**

1. 0.5% Bupivacaine 3 mg/kg
2. epinephrine(1mg/ml) 0.1ml (equal 100mcg)
3. tranexamic acid (500mg/10ml) 1 g
4. ketorolac 30 mg

Locations for injection are quadricep muscle, joint capsule, retropatellar fat pad, medial and lateral collateral ligament

**Intraosseous multimodal drug injection inject the mixture with comprise of**

1. tranexamic acid (500mg/10ml) 1 g
2. ketorolac 30 mg

Location for injection is the intraosseous femoral canal before insert the femoral prothesis

0.5% Bupivacaine 3 mg/kg + epinephrine(1mg/ml) 0.1ml (equal 100mcg) was inject at periarticular location

### Research Design

- A randomized, double-blinded, controlled trial study

### Research Methodology

- **<u>Population and Sample</u>**
- Population: Patients scheduled for elective TKA surgery
- Sample: Patients scheduled for elective TKA surgery at Rajavithi hospital

### Study flowchart

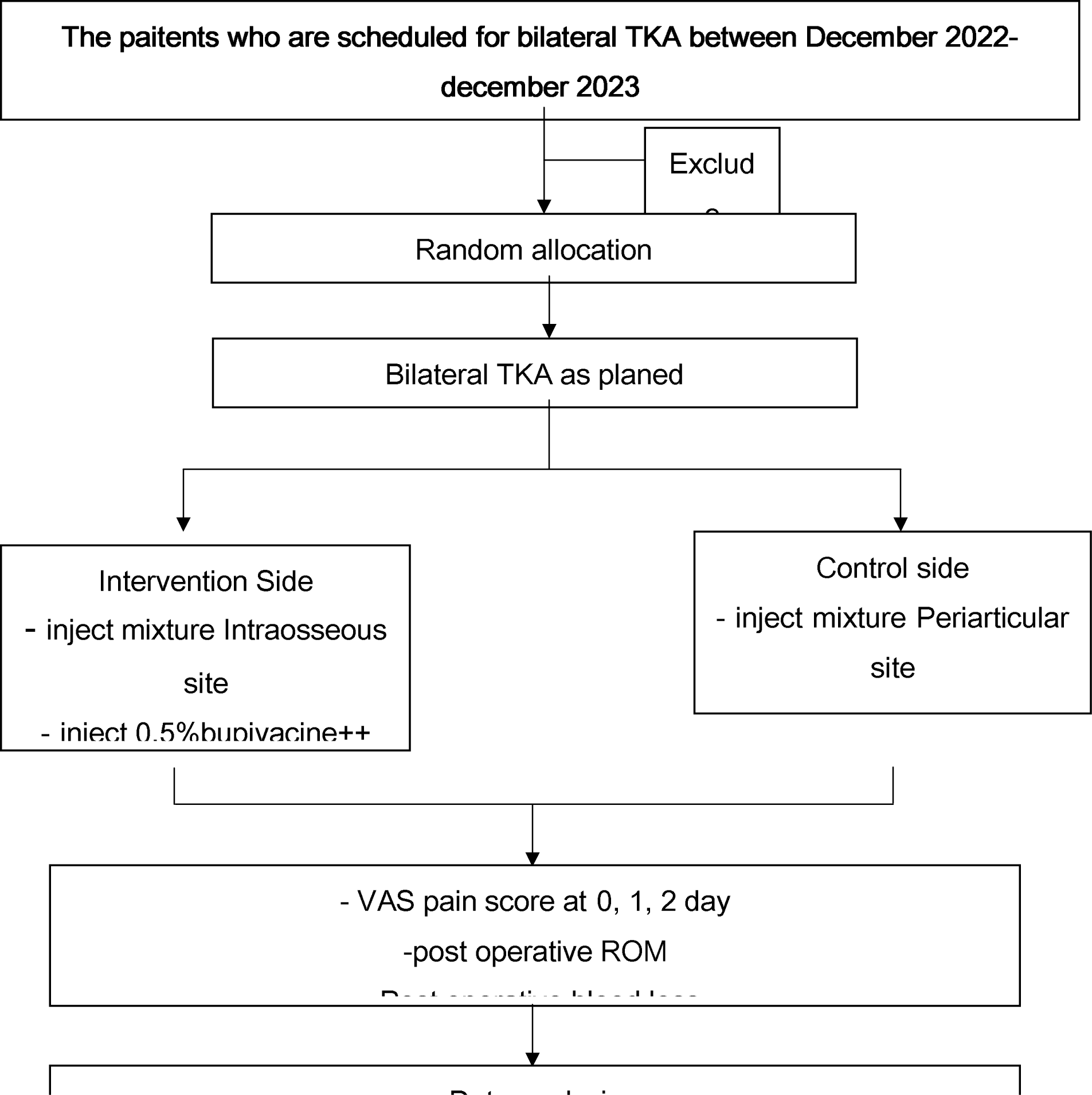

### Inclusion and Exclusion Criteria

- Inclusion Criteria: Primary, bilateral TKA under spinal anesthesia Using PS, cemented design prothesis

-Exclusion criteria: Unilateral TKA Revision TKA History of allergy to intervention medicine Chronic liver and renal failure Pregnancy Previous intraarticular steroid injection Abnormal coagulopathy (INR >1.4 or aPTT ratio > 1.4) Abnormal platelet function or platelet count < 140,0000/mm3 History of DVT Ischemic heart disease Hypertension Arrhythmia

- Withdrawal criteria : Intraoperative complication and postoperative complication Unable to communicate after operation The subject need medication that is interfere with the outcome

- Sample Size Calculation The comparison of the VAS score is the primary determinant of the sample size. The sample size is calculated on MCID (minimal clinically important difference) for VAS score from the study : 1.7 [13], level of significance as 0.05, power as 80%.. Sample size per group is 16.

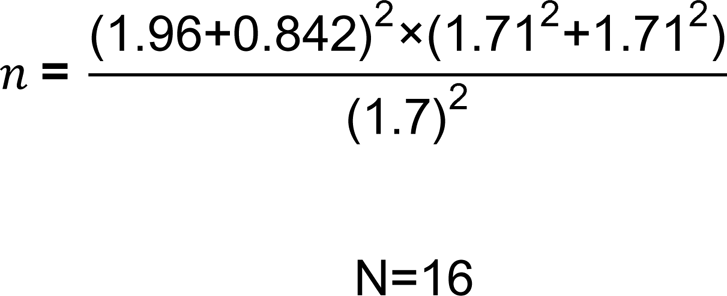

With drop out rate 20% -- N will be 20 for each arm[3]

### Research protocol

-This is a randomized, double-blinded controlled trial study. The study protocol must be approved by the Institutional Review Board, and all subjects must give written informed consent to participate in the study.
- Patients who meet the eligibility criteria are selected to participate in the study. Using a computer-generated block randomization and opaque sealed envelopes, patients are randomly allocated to decide which knee site will be the intervention site or control site. The staff involved in the clinical care, the patients, and assessors are not aware of the treatment assignment.
- Surgical care: Bilateral total knee replacements will be performed using a mid-vastus approach.
  - The experimental intraosseous site: The intraosseous mixture solution will be injected into the medullary canal via the bone-cutting site for preparing to insert a prosthetic joint. The periarticular area will be injected with 0.5% bupivacaine and normal saline (for blinding the surgeon).
  - The control periarticular site: Normal saline will be injected into the medullary canal via the bone-cutting site for preparing to insert a prosthetic joint (for blinding the surgeon). The periarticular area will be injected with the periarticular mixture solution.
-Post-operative care: The postoperative pain management will be given to patients in both groups.
  - NSAID: Naproxen (250) 250 mg oral twice daily.
  - Paracetamol: 500 mg q 4 hrs.
  - Tizanidine: 2 (4 mg) oral q 6 hrs.
  - Gabapentin: 100 mg q 8 hrs.
-The rehabilitation will be scheduled as per protocol.
-VAS pain scores will be recorded at days 0 (6 hours post-operative), 1, and 2 after surgery. The range of motion and postoperative bleeding will also be recorded from the CPM and the drain bottle.

### Outcome Measurements

- Demographic data
- Age, Gender, Weight, Height
- Preoperative pain score
- BMI
- Comorbidity
- severity OA knee score
- Intraoperative data
- Operative time, Anesthesia time
- Blood loss, Intravenous fluid
- Tourniquet, Drain
- Post-operative data

Data Analysis:

- The results were analyzed using SPSS v22.
- P-values and 95% confidence intervals were assessed and reported. A significant P-value is less than 0.05.
- Descriptive statistics were used for the baseline characteristics of the two groups.
- Continuous variables were assessed for normality and presented as the mean and SD or median and range, as appropriate.
- Categorical variables were presented as frequency and percentage.

Between-Group Comparisons:

- The Student’s t-test was used for between-group comparisons of means.
- The Chi-square test was used for between-group comparisons of proportions.
- Two-way repeated measures analysis of variance was used for between-group comparisons of VAS pain score over time.

If the data were repeated measures generalized estimating equations (GEE) and mixed models were used to assess and compare the outcome.

## Risk management protocol

### Severe allergy to the injection Mixture

#### Management

- Excluding patients with known or suspected hypersensitivity to each of the drugs used as a mixture according to the exclusion criteria
- Monitoring for symptoms of side effects from anesthetic drugs throughout the surgery and Postoperative care
- Epinephrine, Dexamethasone, antihistamine are prepared. in the operating room

### Leakage of patient information

#### Management

- The personal data of the research participants will be retained and Individuals data are not publicly disclosed. but The research results will reported as general information. Individual research participants’ data will also be reviewed by the Human Research Ethics Committee of Rajavithi Hospital.

## Data Availability

All data produced in the present study are available upon reasonable request to the authors

